# Deep learning to estimate gestational age from blind ultrasound sweeps of the gravid abdomen

**DOI:** 10.1101/2021.11.22.21265452

**Authors:** Teeranan Pokaprakarn, Juan C. Prieto, Joan T. Price, Margaret P. Kasaro, Ntazana Sindano, Hina R. Shah, Marc Peterson, Mutinta M. Akapelwa, Filson M. Kapilya, Yuri V. Sebastião, William Goodnight, Elizabeth M. Stringer, Bethany L. Freeman, Lina M. Montoya, Benjamin H. Chi, Dwight J. Rouse, Stephen R. Cole, Bellington Vwalika, Michael R. Kosorok, Jeffrey S. A. Stringer

## Abstract

**Background:** Ultrasound is indispensable to gestational age estimation, and thus to quality obstetric care, yet high equipment cost and need for trained sonographers limit its use in low-resource settings.

**Methods:** From September 2018 through June 2021, we recruited 4,695 pregnant volunteers in North Carolina and Zambia and obtained blind ultrasound sweeps (cineloops) of the gravid abdomen alongside standard fetal biometry. We trained a neural network to estimate gestational age from the sweeps and, in three test sets, assessed performance of the model and biometry against previously established gestational age.

**Results:** In our main test set, model mean absolute error (MAE) was 3.9 days (standard error [SE] 0.12) vs. 4.7 days (SE 0.15) for biometry (difference -0.8 days; 95% CI -1.1, -0.5; p<0.001). Results were similar in North Carolina (difference -0.6 days, 95% CI -0.9, -0.2) and Zambia (−1.0 days, 95% CI -1.5, -0.5). Findings were supported in the test set of women who conceived by in vitro fertilization (model MAE 2.8 days [SE 0.28] vs. 3.6 days [SE 0.53] for biometry; difference -0.8 days, 95% CI -1.7, 0.2), and in the set of women from whom sweeps were collected by untrained users with low-cost, battery-powered devices (model MAE 4.9 days [SE 0.29] vs. 5.4 days [SE 0.28] for biometry; difference -0.6, 95% CI -1.3, 0.1).

**Conclusions:** Our model estimated gestational age more accurately from blindly obtained ultrasound sweeps than did trained sonographers performing fetal biometry. These results presage a future where all pregnant people – not just those in rich countries – can access the diagnostic benefits of sonography.

## INTRODUCTION

Accurate estimation of gestational age is fundamental to quality obstetric care. Gestational age is established as early as feasible in pregnancy and then used to determine the timing of subsequent care.^1^ Providers use gestational age to interpret abnormalities of fetal growth, make referral decisions, intervene for fetal benefit, and time delivery. By convention, gestational age is expressed as the time elapsed since the start of the last menstrual period (LMP). Although easily solicited, self-reported LMP has long been recognized as problematic.^2^ Some women may be uncertain of the LMP date. Some (perhaps most^3^) will have a menstrual cycle that varies from the “normal” 28-day length with ovulation on day 14. It is therefore best practice to confirm gestational age dating with an ultrasound exam in early pregnancy. This is achieved by fetal biometry, the measuring of standard fetal structures and applying established formulas.^4^

Although ubiquitous in industrialized regions, obstetric ultrasound is infrequently used in low- and middle-income countries.^5^ Reasons for this disparity include the expense of traditional ultrasound machines, their requirement of reliable electrical power, the need for obstetrics-trained sonographers to obtain images, and the need for expert interpretation. However, two recent developments offer solutions to these obstacles. The first is the availability of low-cost, battery-powered ultrasound devices. There are now more than a dozen manufacturers of low cost probes that can be used with a smart phone or tablet.^6,7^ The second innovation is recent advances in the field of computer vision. Deep learning algorithms are increasingly capable of interpreting radiologic images and these models can be deployed on mobile devices.^8,9^

## METHODS

The Fetal Age Machine Learning Initiative (FAMLI) is an ongoing project that is developing technologies to expand obstetric ultrasound access to low-income settings. Prospective data collection commenced in September 2018 at two sites in Chapel Hill, North Carolina, USA and in January 2019 at two sites in Lusaka, Zambia. We enroll women who are at least 18 years old, have a confirmed single intrauterine pregnancy, and provide written informed consent. The study protocol is approved by the relevant ethical authorities at the University of North Carolina and the University of Zambia.

### Sonography

Each site employs certified sonographers for ultrasound procedures. Participants are recruited during prenatal care and complete a single study visit with no required follow-up; however, we do allow repeat study visits no more frequently than bi-weekly. Evaluation is conducted with a commercial ultrasound machine (multiple makes and models; Table S1). We perform fetal biometry by crown rump length (if <14 weeks) or biparietal diameter, head circumference, abdominal circumference, and femur length (if ≥14 weeks). Each fetal structure is measured twice and the average taken.

During the same examination we also collect a series of blind sweep cineloops. These are free-hand sweeps, approximately 10 seconds in length, across the gravid abdomen in multiple directions and probe configurations. Cranio-caudal sweeps start at the pubis and end at the level of uterine fundus with the probe indicator facing toward the maternal right either perpendicular (90 degrees) or angled (15 and 45 degrees) to the line of probe movement. Lateral sweeps are performed with the probe indicator facing superiorly, starting just above the pubis and sweeping from the left to the right lateral uterine borders and moving cephalad to the uterine fundus. Complete sets of blind sweeps are collected by the study sonographer on both the commercial ultrasound machine and a low-cost, battery-powered device (Butterfly iQ; Guilford, CT, USA). In June 2020, we began collecting a third series of sweeps at the Zambia sites. These “novice blind sweeps” are obtained by a nurse midwife with no training in sonography and include three sweeps in the cranio-caudal axis and three in the lateral axis with the low-cost probe. Prior to obtaining the sweeps, the novice measures the participant’s symphysial-fundal height and sets the depth parameter on the ultrasound device as follows: fundus not palpable = 11cm depth; fundus palpable but <25 cm = 13cm depth; fundus ≥25cm = 15cm depth.

Except for a small number of participants who conceived by *in vitro* fertilization, ground truth gestational age is established by the first ultrasound received. At the North Carolina sites, women present early in pregnancy and gestational age is set according to the American College of Obstetricians and Gynecologists practice guidelines, which incorporate fetal biometry from the first scan and the reported LMP.^4^ At the Zambia sites, women present later in pregnancy^10^ and the LMP is less reliable.^11^ We thus assign gestational age based solely upon the results of the first scan, an approach that antedates the FAMLI protocol.^12,13^

### Training, tuning, and testing datasets

Participants with viable single pregnancies enrolled between September 2018 and June 2021 were included (Figure 1). We applied participant-level exclusions to women whose available medical records did not allow a ground truth gestational age to be established. We applied visit-level exclusions to study scans that (a) did not contain at least two blind sweep cineloops, (b) had missing pixel spacing metadata, or (c) were conducted before 9 weeks of gestation (because they were too infrequent to allow model training). After applying exclusions, we apportioned the remaining data into five non-overlapping groups of participants to develop the deep learning model (training and tuning sets) and evaluate its performance (three test sets).

**Figure 1:**
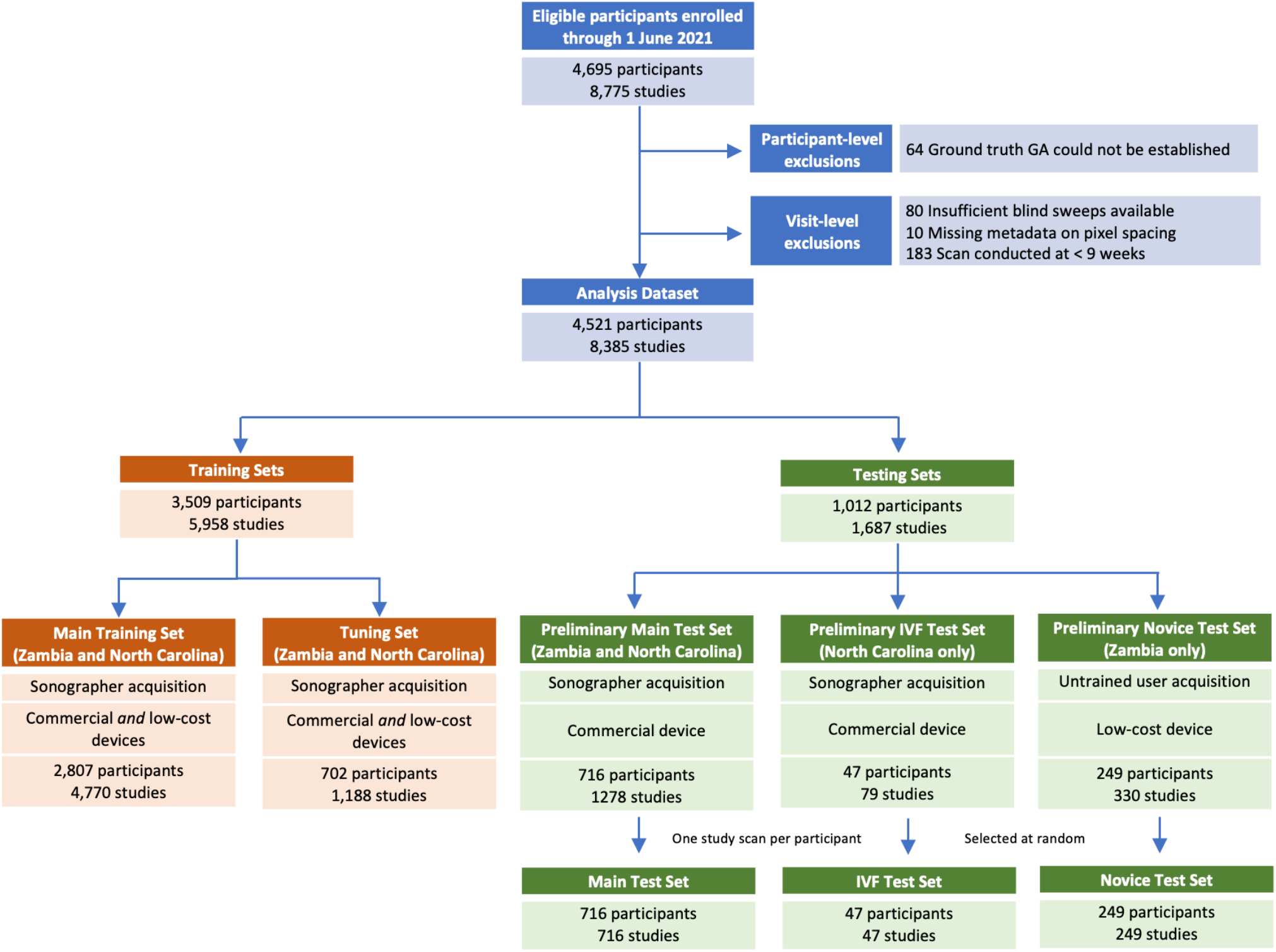
Creation of training and testing datasets After applying participant and visit-level exclusions, we created 2 training sets to develop and tune the deep learning model and 3 test sets to assess its performance. To be eligible for inclusion in a test set a participant must have been dated by a prior scan or in vitro fertilization (IVF). The **IVF test set** comprises all participants who conceived by IVF. The **novice test set** comprises all participants in whom at least one study visit included sweeps collection by a novice user on a low-cost device. The **main test set** was selected at random from among all remaining eligible participants Some participants apportioned to the test sets had contributed more than one study scan; in such cases we selected a single study scan at random. The training sets comprise all participants who remain after creation of the test sets and were split randomly, by participant, in a 4:1 ratio, into a **main training set** and a **tuning set**.

The three test sets were created first. The **IVF test set** comprises women who conceived by *in vitro* fertilization (and thus whose gestational age was known with certainty); all were enrolled in North Carolina. The **novice test set** contains participants who contributed at least one study scan from the novice blind sweep protocol; all were enrolled in Zambia. Our primary assessments are made on an independent **main test set**, which was created as a simple random sample of 30% of eligible women who remained after creation of the other test sets. It includes participants from both Zambia and North Carolina. After establishing the participant members of each test set, we ensured that each woman contributed only a single study scan to her respective test set through random selection (Figure 1). Sensitivity analyses that include all participant study scans are presented in the Appendix.

To be included in a test set, a pregnancy had to be dated by either a prior ultrasound or *in vitro* fertilization; this establishes the ground truth against which both the deep learning model and biometry are measured. In Zambia, a single ultrasound provided by the FAMLI protocol may have been the only scan received. In North Carolina, a single ultrasound provided by the FAMLI protocol may have been conducted on the same day as the participant’s clinical dating ultrasound. In such cases without a prior ground truth benchmark, comparison of the model’s estimate to that of biometry is not possible. These women were thus only included in the datasets used for training. After creation of the three test sets, all remaining participants were randomly allocated in a 4:1 ratio into a **main training set** (80%) and a **tuning set** (20%).

### Technical methods of the deep learning model

Our novel, end-to-end deep learning model takes blind sweep cineloops as input and provides a gestational age estimate as output. Details of the model architecture and constituent parts, including pre-processing steps, training procedure and parameters, and inference procedure are provided in the Appendix.

### Statistical assessment of diagnostic accuracy

Predictive performance of both the model and the biometry is assessed by comparing each approach’s estimate to the previously established ground truth gestational age. The absolute difference between these quantities is the absolute error of the prediction. We report the mean absolute error (MAE) with its standard error (SE), along with the root mean squared error (RMSE) of each approach. We use a paired t-test to assess the mean of the pairwise difference between the model absolute error and the biometry absolute error (|Model Error| - |Biometry Error|). Our null hypothesis was that the mean of this pairwise difference is zero; a negative mean of the pairwise difference whose 95% confidence interval does not include zero would indicate the model to be superior. We compare the model MAE to that of biometry in the overall test datasets and in subsets by geography (Zambia vs North Carolina) and trimester (defined as ≤97 days, 98–195 days, ≥196 days). We also plot the empirical cumulative distribution function (CDF) for the absolute error produced by the model and the biometry. From this empirical CDF, we compare the proportion of study scans in which the absolute error is <7 days and <14 days for the model vs. biometry, using McNemar’s test. Wald-type 95% confidence intervals for the difference in proportions are also computed. Finally, for the novice test set only, we present the diagnostic accuracy of LMP reported at the first patient visit, since this is the relevant comparator for implementation of this technology in low-resource settings.

## RESULTS

From September 2018 through June 2021, 4,695 participants contributed 8,775 ultrasound studies at the four research sites (Figure 1). After applying participant- and visit-level exclusions, we created the three test sets as follows: 712 participants (from both North Carolina and Zambia) formed the main test set; 47 participants (all from North Carolina) formed the IVF test set; 249 participants (all from Zambia) formed the novice test set. As outlined above, participants were allowed to contribute only a single study scan (chosen at random) to their respective test set. The 3,509 participants who remained after creation of the test sets were randomly apportioned into the main training and tuning sets in a 4:1 ratio. Collectively, these women contributed 5,958 study scans containing 109,806 blind sweeps containing 21,264,762 individual image frames for model training and tuning. Baseline characteristics of women included in the combined training sets and the three test sets are presented in Table 1.

**Table 1:** Characteristics of participants in the combined training and tuning sets and in the three test sets

| Characteristic | Training & Tuning Sets<br>N = 3509 | Main Test Set<br>N = 716 | IVF Test Set<br>N = 47 | Novice Test Set<br>N = 249 |
| --- | --- | --- | --- | --- |
| Age, median years (IQR) | 28 (24, 33) | 29 (25, 33) | 37 (33,39) | 27 (23, 30) |
| Body mass index (kg/m<sup>2</sup>)<sup>a</sup> | 24.7 (21.9, 28.5) | 24.8 (22.1, 29.3) | 24.6 (22.7, 28.1) | 24.2 (21.8, 27.2) |
| < 18.5 | 111 (3.1%) | 18 (2.5%) | 1 (2.1%) | 7 (2.8%) |
| 18.5–30 | 2552 (72.7%) | 492 (68.7%) | 34 (72.3%) | 199 (79.9%) |
| > 30 | 617 (17.6%) | 146 (20.4%) | 7 (14.9%) | 22 (8.9%) |
| Parity, median (IQR) | 2 (2, 4) | 2 (1, 4) | 2 (1, 4) | 2 (2, 4) |
| GA at presentation, median weeks (IQR) | 23.0 (13.7, 29.9) | 15.1 (8.8, 24.1) | 9.8 (8.0, 12.3) | 23.7 (19.8, 27.4) |
| No. studies contributed, median (range) | 1 (1, 13) | 2 (1, 11) | 1 (1, 6) | 2 (2, 6) |
| Chronic hypertension, n (%) | 195 (5.6%) | 35 (4.9%) | 0 (0%) | 11 (4.4%) |
| Diabetes, n (%)** | 60 (1.7%) | 23 (3.2%) | 6 (12.8%) | 3 (1.2%) |
| HIV infection, n (%) | 721 (20.5%) | 140 (19.5%) | 0 (0%) | 38 (15.3%) |
| Syphilis in pregnancy, n (%) | 81 (2.3%) | 23 (3.2%) | 0 (0%) | 7 (2.8%) |
| Asthma | 194 (5.5%) | 59 (8.2%) | 8 (17.0%) | 3 (1.2%) |
| Tuberculosis | 80 (2.3%) | 14 (2.0%) | 0 (0%) | 2 (0.8%) |
| Alcohol use during pregnancy | 416 (11.9%) | 146 (20.4%) | 13 (27.7%) | 17 (6.8%) |
| Tobacco use during pregnancy | 74 (2.1%) | 37 (5.2%) | 0 (0%) | 0 (0%) |
| Pregnancy conceived by *in vitro* fertilization | 2 (0.1%) | 0 (0%) | 47 (100%) | 0 (0%) |

<sup>a</sup> Missing height prevents BMI calculation of 5 in IVF test set, 21 in novice test set, 60 in main test set, 230 in combined training sets
<sup>b</sup> Includes both pregestational and “prevalent gestational” (i.e., a participant who would go on to develop gestational diabetes after her last study scan is not counted as diabetic here)

### Model versus biometry in the main test set and IVF test set (Table 2; Figure 2)

In the main test set, the deep learning model outperformed biometry: overall model MAE 3.9 days (SE 0.12) versus biometry MAE 4.7 days (SE 0.15); difference -0.8 days (95% CI: -1.1, -0.5); p<0.001). The observed difference manifested primarily in the third trimester, where the mean of the pairwise difference in absolute error was -1.3 days (95% CI - 1.8, -0.8; p < 0.001). Based on the empirical cumulative distribution function (CDF), the proportion of study scans that were correctly classified within 7 days was higher for the model than for biometry (86.0% vs 77.0%; difference 9.1%; 95% CI: 5.7%, 12.5%; p<0.001). The model similarly outperformed biometry using a 14 day classification window (98.9% vs 96.9%; difference 2.0%; 95% CI: 0.5%, 3.4%; p=0.01).

**Table 2:** Gestational age estimation of deep learning model compared to sonographer in the main test set and IVF test set

|  | Main Test Set (n = 716)<sup>a</sup> |  |  | IVF Test Set (n = 47)<sup>b</sup> |  |  |
| --- | --- | --- | --- | --- | --- | --- |
|  | Model | Biometry | Difference (95% CI) | Model | Biometry | Difference (95% CI) |
| Mean Absolute Error (SE), days | 3.9 (0.12) | 4.7 (0.15) | -0.8 (-1.1, -0.5) | 2.8 (0.28) | 3.6 (0.53) | -0.8 (-1.7, 0.2) |
| Root Mean Square Error, days | 5.1 (0.18) | 6.1 (0.19) | -1.1 (-1.5, -0.6) | 3.4 (0.31) | 5.1 (0.67) | -1.7 (-2.8, -0.5) |
| 1<sup>st</sup> trimester<sup>c</sup> MAE (SE), days | 2.1 (0.19) | 2.0 (0.23) | 0.2 (-0.4, 0.7) | 2.3 (0.34) | 2.2 (0.38) | - |
| 2<sup>nd</sup> trimester<sup>c</sup> MAE (SE), days | 3.1 (0.16) | 3.3 (0.17) | -0.2 (-0.5, 0.1) | 2.7 (0.45) | 3.4 (0.84) | - |
| 3<sup>rd</sup> trimester<sup>c</sup> MAE (SE), days | 4.7 (0.18) | 6.0 (0.22) | -1.3 (-1.8, -0.8) | 3.7 (0.69) | 5.7 (1.38) | - |
| Absolute Error < 7 days (SE), % | 86.0 (1.3) | 77.0 (1.6) | 9.1 (5.7, 12.5) | 95.7 (2.9) | 83.0 (5.5) | - |
| Absolute Error < 14 days (SE), % | 98.9 (0.4) | 96.9 (0.6) | 2.0 (0.5, 3.4) | 100.0 | 100.0 | - |
| North Carolina MAE (SE), days | 3.4 (0.15) | 4.0 (0.20) | -0.6 (-0.9, -0.2) |  |  |  |
| Zambia MAE (SE), days | 4.4 (0.19) | 5.4 (0.22) | -1.0 (-1.5, -0.5) |  |  |  |

<sup>a</sup>The **main test set** comprises a 30% random sample of participants who are dated by a prior ultrasound and who are not included in the IVF or novice test sets; participants enrolled in either North Carolina or Zambia; blind sweeps and fetal biometry were collected by a sonographer on a commercial ultrasound machine. <sup>b</sup>The **IVF test set** comprises all studies conducted in women who conceived by *in vitro* fertilization; all participants were enrolled in North Carolina; blind sweeps and fetal biometry were collected by a sonographer on a commercial ultrasound machine. <sup>c</sup>Trimesters defined as ≤97 days, 98 – 195 days, ≥196 days; MAE = mean absolute error

**Figure 2:**
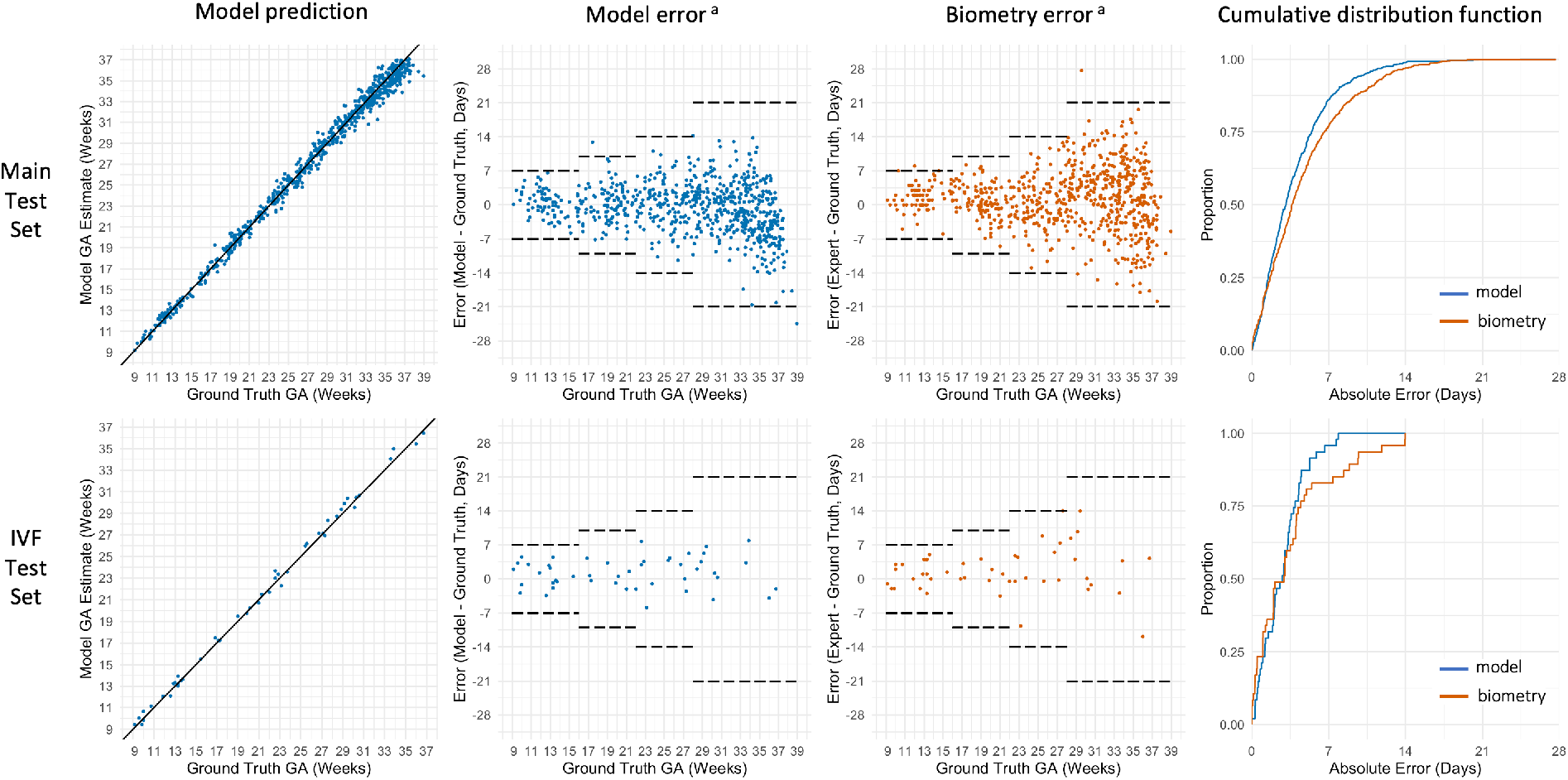
Gestational age estimation of deep learning model compared to trained sonographer in the main and IVF test sets ^a^ dashed horizontal lines represent expected accuracy of ultrasound biometry^4^

Among the 47 study scans in the IVF test set, the model MAE was 2.8 days (SE 0.28) compared to MAE of 3.6 days (SE 0.53) for biometry (difference = -0.8 days; 95%CI: -1.7, 0.2; p = 0.10). As was observed in the main test set, the difference was most pronounced in the third trimester where the estimated mean of the pairwise difference in absolute error was -2.0 days. Based on the empirical CDF, the proportion of study scans that were correctly classified within 7 days was higher for the model than for biometry (95.7% vs 83.0%).

Owing to the small sample size in our IVF test set, we did not perform statistical tests on the difference by trimester or the difference in proportion. Both model and biometry correctly categorized 100% of cases within 14 days (Table 2).

### Model versus biometry and LMP in the novice test set (Table 3; Figure 3)

The novice test set contains 249 sets of blind sweeps obtained on a low-cost, battery-powered device by an untrained user. As above, we compared model estimates to biometry obtained by a trained sonographer on a commercial ultrasound. But we also compared the model estimates to the gestational age that would have been calculated had only the LMP been available (as is overwhelmingly the case in Zambia). In the novice test set, the model and biometry performed similarly: overall model MAE 4.9 days (SE 0.29) versus biometry MAE 5.4 days (SE 0.28); difference -0.6 days (95% CI: 1.3, 0.1); p=0.11). However, when compared to LMP, the model was clearly superior: model MAE 4.9 days (SE 0.29) versus LMP MAE 17.4 days (SE 1.17); difference -12.7 days (95% CI: -15.0, -10.3); p<0.001). Based on the empirical CDF, the proportion of study scans that were correctly classified within 7 days was substantially higher for the model than for LMP (71.9% vs 40.1%; difference 36.1% [95% CI 28.0%, 44.2%]; p<0.001). The model similarly outperformed LMP using a 14-day classification window (94.8% vs 55.1%; difference 40.5% [95% CI 33.9%, 47.1%]; p<0.001).

**Table 3:**
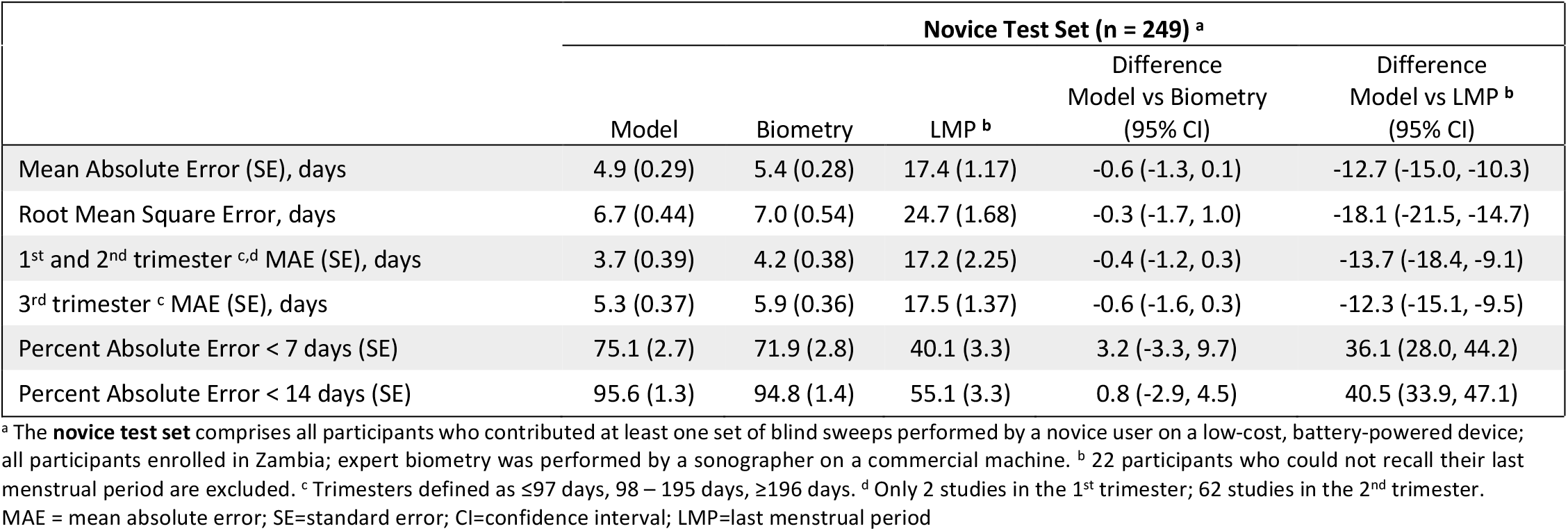
Gestational age estimation of deep learning model compared to trained sonographer in the novice test set

**Figure 3:**
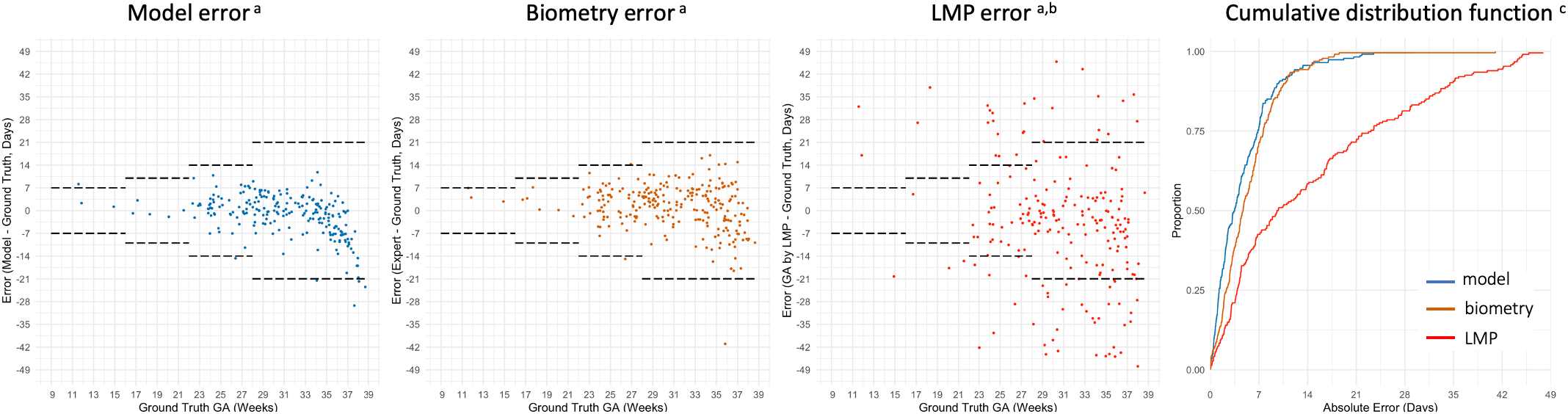
Gestational age estimation of deep learning model compared to trained sonographer and last menstrual period (LMP) in the novice test set ^a^ dashed horizontal lines represent expected accuracy of ultrasound biometry^4^ ^b^ data missing from 22 participants who could not recall their LMP ^c^ 13 studies from GA by LMP excluded from the plot because the absolute error is truncated at 49 days

## DISCUSSION

Quality obstetric care requires accurate knowledge of gestational age. We built a deep learning model that can perform this critical assessment from blindly obtained ultrasound sweeps of the gravid abdomen. Expressed as mean absolute error or as the proportion of estimates that falls within 7 or 14 days of a previously defined ground truth gestational age, the model performance is superior to that of a trained sonographer performing fetal biometry on the same day. Results were consistent across geographical sites and supported in a test set of women who conceived by IVF (whose gestational age is unequivical) and in a test set of women from whom the ultrasound blind sweeps were obtained by a novice provider using a low-cost, battery-powered device.

This research addresses a critical shortcoming in the delivery of obstetrical care in low- and middle-income countries. The Lusaka public sector is typical of care systems across the African sub-continent and parts of Asia in that few women have access to ultrasound pregnancy dating and the median gestational age at presentation is 23 weeks (IQR 19, 26).^10^ This means that each year in the city of Lusaka, more than 100,000 pregnancies^14^ must be managed with an unacceptably low level of gestational age precision (Figure 3).^11,15^ The availability of a resource-appropriate technology that could accurately assign gestational age in the late second and third trimesters could be transformative in Lusaka and similar settings around the world.

Recent years have seen an explosion in the application of deep learning to healthcare, particularly the interpretation of medical images.^9^ Successful projects to date have leveraged extant clinical datasets and trained machine learning algorithms on ideally-obtained single images (e.g., ocular fundus,^16,17^ breast,^18^ skin^19^) that have been annotated by human experts. In contrast, our study collects thousands of images from each participant in the form of blind sweeps. Each cineloop frame in the sweep is itself a two-dimensional ultrasound image that is provided to the neural network during training. Although most of these frames would be considered clinically sub-optimal views, the sheer number of them (more than 21 million) provides a comprehensive picture of the developing fetus from every conceivable angle throughout gestation. Considered as individual images rather than participants or studies or sweeps, our training set is two orders of magnitude larger than most of the prior high-profile applications of deep learning to medical imaging.^16-19^ This may explain why the model so consistently outperforms expert biometry even when some of our training data include studies from women who present late for care and whose clinically established gestational age may be subject to measurement error.

Strengths of this study include its prospective nature, bespoke blind sweep sonography procedures, and inclusion of women from both high- and low-resource health care systems. We used several different makes and models of ultrasound scanners for data collection, a feature that likely bolsters the model’s generalizability. Although we did not deliberately impose a lower gestational age limit on enrollment, our dataset includes very few scans at < 9 gestational weeks and we thus are unable to make estimates below this threshold. Data were similarly sparse beyond 37 weeks (term gestation) and the model appears to systematically underestimate gestational age beyond this point in the novice test set. We note however that this limitation seems likely to affect only a minority of women – those who seek prenatal care but who have no visits between 9 and 37 weeks. From our prior population-based study of 115,552 pregnancies in Lusaka, under 1% of women would meet these criteria.^10^ Finally, we acknowledge that our blind sweep approach would be a sea change, and might be seen as a threat to obstetric ultrasound capacity building in low-resource settings. Nonetheless, our data suggest that at least for gestational age estimation, capacity building efforts may be better directed elsewhere, as our approach is robust, and because it would be made freely available, affordable and scalable.

Beyond gestational age determination, obstetrical ultrasound can diagnose a wide range of conditions that may result in preventable morbidity or death, including ectopic pregnancy, multiple gestation, placenta previa, fetal demise, growth restriction, disorders of amniotic fluid volume, abnormal fetal blood flow, malpresentation, and fetal anatomic anomalies. Whether any of these diagnoses would be amenable to a deep learning approach is unknown but ongoing vibrant research at the intersection of artificial intellegence and obstetric sonography is promising.^20,21^ Given the ever-expanding computational capacity of mobile devices and the real advances that have been made in low-cost sonography, it seems only a matter of time until the world’s most remote and under-resourced obstetrical services have access to the full diagnostic power of ultrasound.

## Data Availability

All data produced in the present study are available upon reasonable request to the authors through a third party data sharing mechanism and subject to specific terms around attribution.

## Ethical Appovals

This study protocol was approved by the University of North Carolina Institutional Review Board, the University of Zambia Biomedical Research Ethics Committee, and the Zambia National Health Research Authority prior to initiation.

## Works Cited

1. World Health Organization. WHO recommendations on antenatal care for a positive pregnancy experience (2016). Available at: https://www.who.int/reproductivehealth/publications/maternal_perinatal_health/anc-positive-pregnancy-experience/en/ Accessed 18 November 2021.

2. Kramer MS, McLean FH, Boyd ME, Usher RH. The validity of gestational age estimation by menstrual dating in term, preterm, and postterm gestations. Jama. 1988;260(22):3306–3308.

3. Matsumoto S, Nogami Y, Ohkuri S. Statistical studies on menstruation: a criticism on the definition of normal menstruation. Gunma J Med Sci 1962;11:294–318.

4. American College of Obstetricians and Gynecologists, American Institute of Ultrasound in Medicine, Society for Maternal-Fetal Medicine. Committee Opinion No 700: Methods for Estimating the Due Date. Obstetrics and gynecology. 2017;129(5):e150–e154.

5. Yadav H, Shah D, Sayed S, Horton S, Schroeder LF. Availability of essential diagnostics in ten low-income and middle-income countries: results from national health facility surveys. The Lancet Global health. 2021.

6. Marsh-Feiley G, Eadie L, Wilson P. Telesonography in emergency medicine: A systematic review. PloS one. 2018;13(5):e0194840.

7. Becker DM, Tafoya CA, Becker SL, Kruger GH, Tafoya MJ, Becker TK. The use of portable ultrasound devices in low-and middle-income countries: a systematic review of the literature. Trop Med Int Health. 2016;21(3):294–311.

8. Carin L, Pencina MJ. On Deep Learning for Medical Image Analysis. In: Livingston EH, Lewis RJ, eds. JAMA Guide to Statistics and Methods. New York, NY: McGraw-Hill Education; 2019.

9. Esteva A, Robicquet A, Ramsundar B, et al. A guide to deep learning in healthcare. Nature medicine. 2019;25(1):24–29.

10. Chi BH, Vwalika B, Killam WP, et al. Implementation of the Zambia electronic perinatal record system for comprehensive prenatal and delivery care. International journal of gynaecology and obstetrics: the official organ of the International Federation of Gynaecology and Obstetrics. 2011;113(2):131–136.

11. Price JT, Winston J, Vwalika B, et al. Quantifying bias between reported last menstrual period and ultrasonography estimates of gestational age in Lusaka, Zambia. International journal of gynaecology and obstetrics: the official organ of the International Federation of Gynaecology and Obstetrics. 2019;144(1):9–15.

12. Castillo MC, Fuseini NM, Rittenhouse K, et al. The Zambian Preterm Birth Prevention Study (ZAPPS): Cohort characteristics at enrollment. Gates Open Res. 2018;2:25.

13. Price JT, Vwalika B, Rittenhouse KJ, et al. Adverse birth outcomes and their clinical phenotypes in an urban Zambian cohort. Gates Open Res. 2019;3:1533.

14. Zambian Ministry of Health. Annual Health Statistics Report 2017-2019; Available at https://www.moh.gov.zm/?wpfb_dl=159 (accessed 18 November 2021). October 2020, Lusaka.

15. Vwalika B, Price JT, Rosenbaum A, Stringer JSA. Reducing the global burden of preterm births. The Lancet Global health. 2019;7(4):e415.

16. Gulshan V, Peng L, Coram M, et al. Development and Validation of a Deep Learning Algorithm for Detection of Diabetic Retinopathy in Retinal Fundus Photographs. Jama. 2016;316(22):2402–2410.

17. Milea D, Najjar RP, Zhubo J, et al. Artificial Intelligence to Detect Papilledema from Ocular Fundus Photographs. The New England journal of medicine. 2020;382(18):1687–1695.

18. McKinney SM, Sieniek M, Godbole V, et al. International evaluation of an AI system for breast cancer screening. Nature. 2020;577(7788):89–94.

19. Esteva A, Kuprel B, Novoa RA, et al. Dermatologist-level classification of skin cancer with deep neural networks. Nature. 2017;542(7639):115–118.

20. Maraci MA, Yaqub M, Craik R, et al. Toward point-of-care ultrasound estimation of fetal gestational age from the trans-cerebellar diameter using CNN-based ultrasound image analysis. J Med Imaging (Bellingham). 2020;7(1):014501.

21. van den Heuvel TLA, Petros H, Santini S, de Korte CL, van Ginneken B. Automated Fetal Head Detection and Circumference Estimation from Free-Hand Ultrasound Sweeps Using Deep Learning in Resource-Limited Countries. Ultrasound Med Biol. 2019;45(3):773–785.

