## Appendix for "Deep learning to estimate gestational age from blind ultrasound sweeps of the gravid abdomen"

#### Section 1: Technical Methods for the Deep Learning Model

**This section is redacted until the manuscript is accepted for publication**

#### Section 2: Additional Statistical Methods

##### Calculation of Standard Errors (SE) for Root Mean Square Error (RMSE)

Let $U_i, i = 1, \ldots, n$ be $n$ independent errors.
Denote RMSE as $\hat{\sigma}_n$, where $\hat{\sigma}_n^2 = n^{-1} \sum_{i=1}^{n} U_i^2$.
Define

$$\hat{\tau}_n^2 = \frac{\sum_{i=1}^{n}\left[U_i^4 - \hat{\sigma}_n^4\right]}{(4n\hat{\sigma}_n^2)}$$

Then the SE for the RMSE is $\hat{\tau}_n/\sqrt{n}$ based on the delta method.

##### Calculation of Standard Errors (SE) and Confidence Interval for Difference in Root Mean Square Error (RMSE)

Let $(X_1, Y_1), \ldots, (X_n, Y_n)$ be the errors for the model ($X$) and for the expert (Y). Let $\hat{\sigma}_n^2 = \mathrm{P}_n X^2$ and $\hat{\tau}_n^2 = \mathrm{P}_n Y^2$, where $\mathrm{P}_n$ is the empirical process (i.e., $\mathrm{P}_n f(X) = n^{-1} \sum_{i=1}^{n} f\,(X_i)$). Also, denote $\sigma_0^2 = PX^2$ and $\tau_0^2 = PY^2$, where $P$ is the expectation.

We can use the Taylor expansion, and the fact that the derivative of $\sqrt{u}$ is $u^{-1/2}/2$, to obtain that $\sqrt{n}\left(\sqrt{\hat{\sigma}_n^2} - \sigma_0\right) = \sqrt{n}(\hat{\sigma}_n^2 - \sigma_0^2)/(2\sigma_0) + o_P(1)$. Similarly, we can verify that $\sqrt{n}\left(\sqrt{\hat{\tau}_n^2} - \tau_0\right) = \sqrt{n}(\hat{\tau}_n^2 - \tau_0^2)/(2\tau_0) + o_P(1)$. Now, letting $IF(X, Y) = (X^2 - \sigma_0^2)/(2\sigma_0) - (Y^2 - \tau_0^2)/(2\tau_0)$, we have that $D_n = \sqrt{\hat{\sigma}_n^2} - \sqrt{\hat{\tau}_n^2} - \sigma_0 + \tau_0 = n^{-1/2}\mathrm{P}_n IF(X, Y) + o_P\left(n^{-1/2}\right)$.
This means that the true variance of $D_n$ equals $n^{-1}P\left(IF(X, Y)\right)^2 + o(1)$.

Let $\widehat{IF}(X, Y) = (X^2 - \hat{\sigma}_n^2)/(2\hat{\sigma}_n) - (Y^2 - \hat{\tau}_n^2)/(2\hat{\tau}_n)$.
$n^{-1} \sum_{i=1}^{n}\left[\widehat{IF}(X_i, Y_i)\right]^2$ is consistent for $P[IF(X, Y)]^2$, and thus we can consistently estimate the the SE of the difference between the RMSEs with

$$n^{-1/2}\sqrt{n^{-1}\sum_{i=1}^{n}\left[\widehat{IF}(X_i, Y_i)\right]^2}$$

### Section 3: Supplementary Figures

#### Figure S1: DL model architecture

**This Figure is redacted until the manuscript is accepted for publication**

#### Figure S2: Gestational age distribution of the training and testing sets

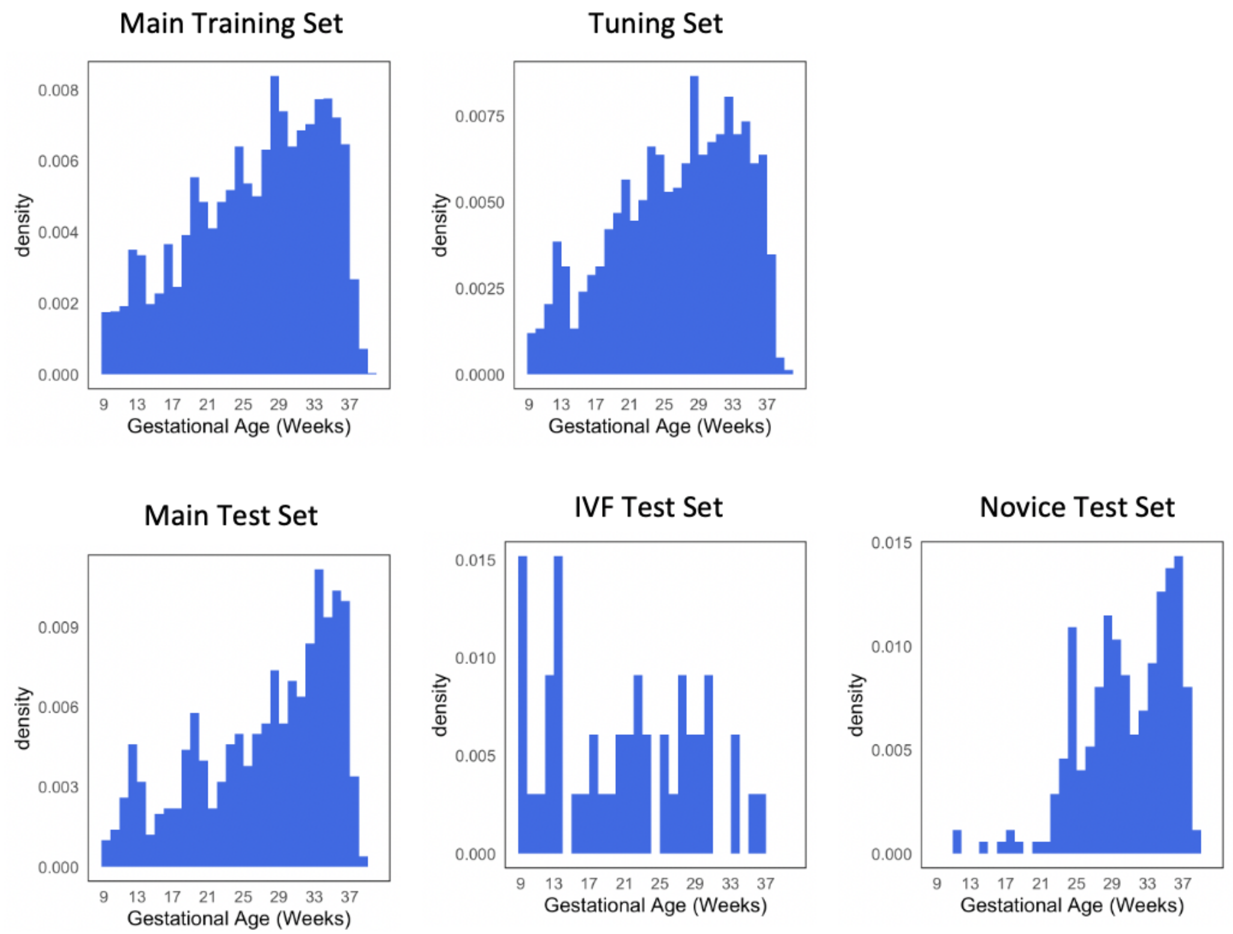

### Figure S3: Overview of FAMLI protocol clinical data collection

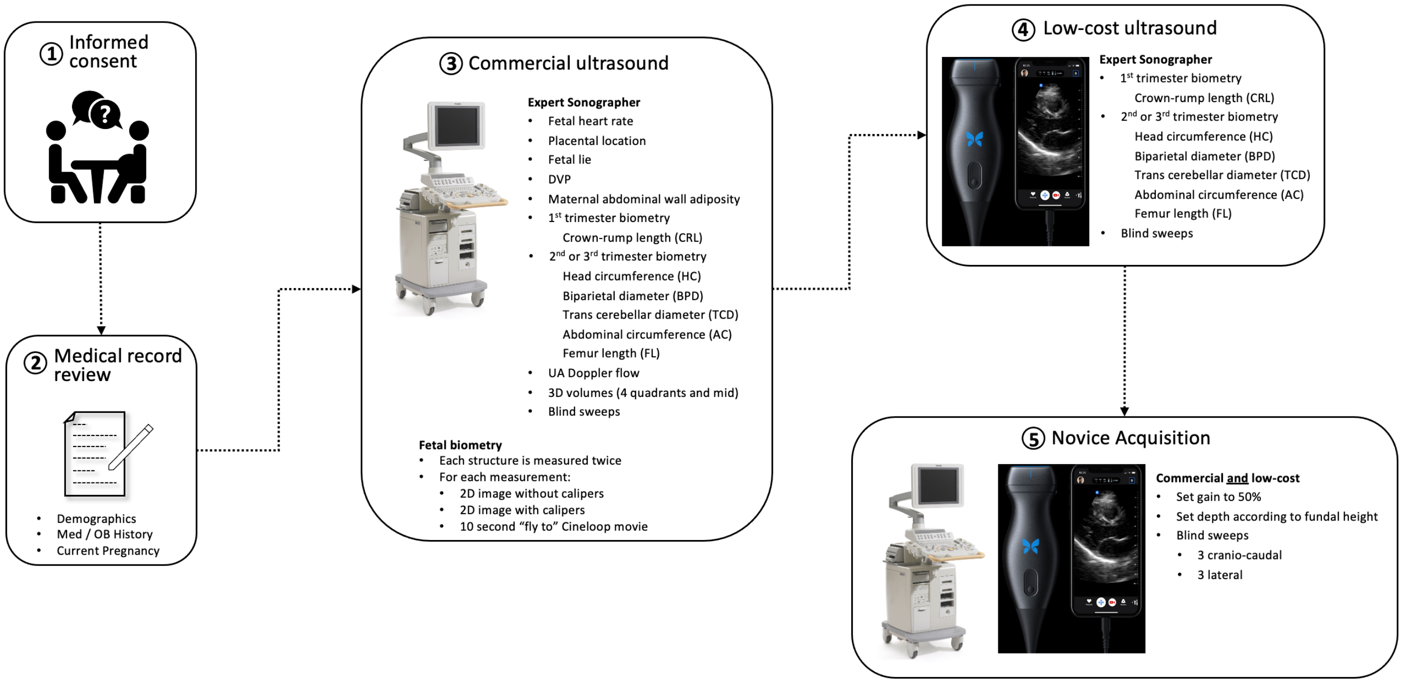


Graphical representation of a participant visit and ultrasound data collection in the FAMLI Study. Step 5 (novice acquisition) began in June 2020 at the Zambia sites only.

### Section 4: Supplemental Tables

**Table S1:** Ultrasound devices used

| Ultrasound Make and Model | Training Set N=4,770 | Tuning Set N=1,188 | Main Test Set N=716 | IVF Test Set N=47 | Novice Test Set N=249 |
|---|---|---|---|---|---|
| Butterfly iQ | 3833 | 941 | 0 | 0 | 249 |
| GE LOGIQ C3 Premium | 83 | 17 | 0 | 0 | 0 |
| GE LOGIQ e | 136 | 35 | 0 | 0 | 0 |
| Sonosite MTurbo | 1955 | 457 | 219 | 0 | 0 |
| GE Voluson E8 | 801 | 236 | 188 | 17 | 0 |
| GE Voluson S6 | 1712 | 417 | 299 | 30 | 0 |

Each participant study visit involves data collection with both a commercial and low-cost device. We limited the test sets to a single device (Main test set and IVF test set has commercial device only; Novice test set has low-cost device only). We did not impose this limitation on the training and tuning sets (i.e., during training a single participant study could contribute blind sweep cineloops from two devices.) Butterfly = Butterfly Network, Inc Guilford, CT, USA; GE = General Electric Healthcare, Zipf, Austria; Sonosite = SonoSite Inc, Bothell, WA, USA.

**Table S2:** Gestational age estimation of deep learning model compared to expert sonographer – sensitivity analysis

| | **Main Test Set (n = 1278)[a]** | | | **IVF Test Set (n = 79)[b]** | | |
|---|---|---|---|---|---|---|
| | Model | Biometry | Difference (95% CI) | Model | Biometry | Difference (95% CI) |
| Mean Absolute Error (SE), days | 3.9 (0.09) | 4.7 (0.11) | -0.8 (-1.1, -0.6) | 3.0 (0.26) | 3.6 (0.38) | -0.6 (-1.3, 0.1) |
| Root Mean Square Error, days | 5.1 (0.13) | 6.2 (0.15) | -1.1 (-1.4, -0.8) | 3.8 (0.32) | 4.9 (0.47) | -1.1 (-2.0, -0.2) |
| 1st trimester [c] Mean Absolute Error (SE), days | 2.3 (0.17) | 2.2 (0.18) | 0.1 (-0.4, 0.5) | 2.2 (0.34) | 2.4 (0.43) | - |
| 2nd trimester Mean Absolute Error (SE), days | 3.1 (0.11) | 3.5 (0.13) | -0.4 (-0.7, -0.2) | 2.5 (0.30) | 3.0 (0.54) | - |
| 3rd trimester Mean Absolute Error (SE), days | 4.8 (0.15) | 6.1 (0.18) | -1.3 (-1.7, -0.9) | 4.2 (0.58) | 5.1 (0.76) | - |
| Absolute Error < 7 days (SE), % | 85.4 (1.0) | 77.3 (1.2) | 8.1 (5.6, 10.7) | 92.4 (3.0) | 84.8 (4.0) | - |
| Absolute Error < 14 days (SE), % | 98.7 (0.3) | 96.5 (0.5) | 2.3 (1.1, 3.4) | 100.0 | 100.0 | - |
| North Carolina Mean Absolute Error (SE), days | 3.6 (0.12) | 4.1 (0.14) | -0.5 (-0.7, -0.2) | - | - | - |
| Zambia Mean Absolute Error (SE), days | 4.2 (0.15) | 5.5 (0.18) | -1.3 (-1.7, -0.9) | - | - | - |

Our primary analyses limited test sets to a single ultrasound study per participant. This sensitivity analysis allows participants to contribute more than one study to their test set. [a] The **main test set** comprises a 30% random sample of participants who are dated by a prior ultrasound and who are not included in the IVF or novice test sets; participants enrolled in either North Carolina or Zambia; blind sweeps and fetal biometry were collected by a sonographer on a commercial ultrasound machine. [b] The **IVF test set** comprises all studies conducted in women who conceived by *in vitro* fertilization; all participants were enrolled in North Carolina; blind sweeps and fetal biometry were collected by a sonographer on a commercial ultrasound machine. [c] Trimesters defined as ≤97 days, 98 – 195 days, ≥196 days. SE=standard error; CI=confidence interval; LMP=last menstrual period

**Table S3:** Gestational age estimation of deep learning model compared to expert sonographer – sensitivity analysis

| | Novice Test Set (n = 330) [a] | | | | |
|---|---|---|---|---|---|
| | Model | Biometry | LMP [b] | Difference Model vs Expert (95% CI) | Difference Model vs LMP[b] (95% CI) |
| Mean Absolute Error (SE), days | 5.0 (0.27) | 5.5 (0.26) | 17.9 (1.06) | -0.5 (-1.1, 0.1) | -13.1 (-15.2, -11.0) |
| Root Mean Square Error, days | 7.0 (0.41) | 7.3 (0.49) | 25.6 (1.55) | -0.3 (-1.3, 0.8) | -18.8 (-21.9, -15.6) |
| 1st and 2nd trimester [c,d] Mean Absolute Error (SE), days | 3.9 (0.37) | 3.9 (0.33) | 16.5 (1.90) | -0.0 (-0.7, 0.7) | -13.1 (-17.0, -9.2) |
| 3rd trimester Mean Absolute Error (SE), days | 5.4 (0.34) | 6.1 (0.33) | 18.4 (1.28) | -0.7 (-1.5, 0.1) | -13.1 (-15.6, -10.6) |
| Absolute Error < 7 days (SE), % | 74.5 (2.4) | 70.9 (2.5) | 39.5 (2.8) | 3.6 (-2.2, 9.5) | 37.2 (30.1, 44.2) |
| Absolute Error < 14 days (SE), % | 93.9 (1.3) | 94.2 (1.3) | 54.7 (2.9) | -0.3 (-3.7, 3.1) | 39.9 (34.0, 45.7) |

Our primary analyses limited test sets to a single ultrasound study per participant. This sensitivity analysis allows participants to contribute more than one study to their test set. [a] The **novice test set** comprises all participants who contributed at least one set of blind sweeps performed by a novice user on a low-cost, battery-powered device; all participants enrolled in Zambia; expert biometry was performed by a sonographer on a commercial machine. [b] 22 participants who could not recall their last menstrual period are excluded. [c] Trimesters defined as ≤97 days, 98 – 195 days, ≥196 days. [d] Only 2 studies in the 1st trimester; 62 studies in the 2nd trimester. SE=standard error; CI=confidence interval; LMP=last menstrual period.
